# Challenges Faced by Dialysis Unit Staff during COVID −19 times-A Qualitative Study

**DOI:** 10.1101/2020.10.20.20215715

**Authors:** P. RaviKumar, Amol Dongre

## Abstract

**Introduction:** The novel SARS COV2-Covid −19 has become a global pandemic since January 2020 and has been spreading exponentially. Dialysis patients with lowered immunity are at high risk. The dialysis patients come for repeated treatment. Hence the dialysis staffs are also at great risk of contracting COVID-19.

**Objective:** To study the challenges faced by the dialysis staff during the COVID-19 pandemic in a rural hemodialysis unit.

**Material and Methods:** The study was conducted in the hemodialysis unit of Sri ManakulaVinayagar Medical College Hospital, Puducherry, India. We did free list and pile sorting to understand the salient problems and its structure as perceived by the team members. We found Smith’s S value for free list. Multidimensional scaling and Hierarchical cluster analysis were done to pile sort data. Data was analyzed using Anthropac 4.983/X. In addition, group interview was done to get in-depth information and validate the findings obtained from the free list and pile sort exercise.

**Results:** Twelve salient items were generated from the free list. In pile sort, we got three broad domains-the shortage of personal safety equipment, the lack of personal safety and presence of logistical and operational problems. Relative to other items, testing by RT-PCR was surprisingly not perceived to be important for them.

**Conclusion:** Addressing the shortage of personal safety equipment, impediments to personal safety and giving credence to the feelings, fears and needs of the dialysis staff in a dialysis unit during COVID −19 pandemic are paramount in ensuring their safety and improving working dynamics.

## Introduction

The COVID-19 pandemic sweeping across the whole globe has affected everyone in diverse ways. The Chronic kidney disease patients have been affected in a very big way as they continue to visit the hospitals and hemodialysis units for continuity of care and to get regular dialysis sessions. The dialysis patients visit their dialysis centre at least twice or traditionally thrice a week. The dialysis staffs have also been working hard during these challenging times. Health care workers in the United States have expressed concerns about working with patients during the active pandemic.^1^ These challenges are complex and interact with each other. Hence, to prioritize the intervention and understand its complexity and interaction, we did a qualitative evaluation to understand the problem from team members’ point of view. The fact of the matter is though qualitative research is increasingly used in many health care fields it is underused and rarely reported in Nephrology high impact journals.^2^

### Objective

The objective of the present study was to understand the challenges faced by the staff working in the times of COVID −19 pandemic in the hemodialysis unit in a rural medical setting.

## Methods

### Setting

The setting of the data collection and the study was in the main dialysis unit of Sri ManakulaVinayagar Medical College and Hospital, Madagadipet, Puducherry 605107, India. Accreditation of the unit: This dialysis unit has been audited on a regular basis by an external Nephrology consultant auditor since last six years. It has also passed the step ½ of the NABH accreditation process.

### Design

It was an exploratory type of qualitative research using free list and pile sort exercise. Free listing is an exercise which involves collecting mental thoughts in a dimension. It depicts the cultural salience of these thoughts within these groups. Written free listing done with qualified staff actually helps in rapid data collection.^3,4^ It helps in identifying items in a cultural domain, indicates, which of the things are most relevant and shows us the extent of variation regarding the beliefs being probed,^3,4,5^ Pile sorting is also an exercise which tell us how people think about certain ideas and how they organize their thoughts, about how they value things and attach importance to some particular themes.

### Sample size and Sampling

Twelve dialysis unit staff members were interviewed as part of a FGD in the main hemodialysis unit of the Sri Manakula Vinayagar Medical College and Hospital. This is a busy hemodialysis unit catering to around 1300-1500 dialysis sessions a month. The patients have 24-hour access to dialysis facilities.

### Data collection and analysis method

After exchanging pleasantries in the group and breaking the ice, “each dialysis unit staff was individually asked to make the free list of challenges faced by them during their work in the dialysis unit in these COVID-19 days. Each one of them wrote their responses on a paper. Participants were asked a primary stimulus question – “*Please write down as many challenges as you face while working in dialysis unit during this Covid time*”. The responses for each of the participant was entered in notepad and analyzed using the Visual Anthropac software package. The Smith’s S value was calculated to identify the more prominent item in all the lists. We analyzed free listing using Anthropac software to get the most salient challenges faced by the team.

To understand the structure of the domain, we selected top 12 salient items for pile sorting. The point at which the Smith’s S score showed a sharp decline was taken a cut-off. All 12 participants were invited to join the pile sort exercise. Each of them was individually asked to group these items together to form the piles according to their criteria. Each staff member was interviewed again separately and the reasons for their individual groupings were gathered. After pile sorting was done, a multi-dimensional scaling and hierarchical cluster analysis was done using Anthropac 4.983/X. Hierarchal mode of digital visualization of the perceptions in a cognitive domain emerged from the software. The concept webbing or mental tapping^5^ done here was a dynamic group interaction in the dialysis unit. The identification of the group in study was dialysis nurses, technicians, and doctors. The key question in focus was their challenges during the present day COVID-19 pandemic. After the free listing and pile sorting was carried out, the input available from the listing and piles were taken up for analysis. Here the major statistical analyses done were done by a method called as multidimensional scaling where all the pooled data (statements) was put into a basic map by using computer software. Here each description/labels/statement was represented by a point on the map. The main analysis involved taking the statements and dividing the map into groups or clusters,^,6,7,8,9^ which should be logical and interpretative. After the concepts or labels were analyzed and interpreted, the results were available for further use. The final stage involved using the main thrust of the thoughts and ideas to be visualized clearly on the map generated as a form of pictorial depiction.^10,11^ so as to bolster new changes or wants to be addressed for giving better care to our patients during these stressful and challenging times.

## Results

Out of 12 respondents, five were nurses, five were technicians and two were doctors. Among them, three were males and the remaining nine were females. Table-1 shows free listing of twelve salient items with descending order of salience. As seen in Supplemental S Figure-1, the structure of the cognitive domain of challenges faced by staff consisted of three sub-groups – 1) operational problems, 2) the major group consisted of items related to lack of personal safety for staff and 3) concerns for lack of personal safety equipment for the staff. Supplemental S Figure 2 shows the Algorithm (Flow chart)followed in the study.

**Table-1:** Free list of salient items (challenges) faced by the dialysis unit staff.

| **Item** | **Frequency (%)** | **Average Rank** | **Salience** |
| --- | --- | --- | --- |
| Lack of PPEs | 75 | 3.56 | 0.607 |
| Patient's lacking public transport | 75 | 5 | 0.523 |
| Lack of sanitizer / soap solutions | 83.3 | 6 | 0.505 |
| Difficulty in scheduling of patients | 66.7 | 4.75 | 0.482 |
| Incorrect history given by patients | 66.7 | 5.38 | 0.436 |
| Not testing for COVID-19 RT-PCR presently | 41.7 | 4 | 0.29 |
| Possible Covid positivity in patient | 33.3 | 3 | 0.286 |
| Hot water unavailable | 50 | 6.17 | 0.278 |
| Relatives do not follow isolation rules | 66.7 | 9.25 | 0.249 |
| Uncooperative patient/relative wear no masks | 66.7 | 9.75 | 0.24 |
| Financial stringency | 41.7 | 7.8 | 0.213 |
| Face mask shortage | 25 | 4.33 | 0.199 |

## Discussion

The main finding as shown in figure 1, was about the issue of shortage of Personal safety equipment (i.e. Lack of sanitizer, soap solutions, lack of hot water in the unit, lack of PPEs, face mask shortage) which stood apart closely together despite repeated modeling a number of times on the software of the computer. The second finding as a cluster was that of the danger to personal safety of the staff (Incorrect history given by patients, relatives not following isolation rules, patients and relatives not wearing masks, unavailability of testing for RT-PCR, fear/Possibility of Covid-19 positivity. The third finding as a cluster highlighted was that of the logistics and operational issues (Patient lacking public transport, difficulty in scheduling of patients, financial stringency)

**Figure 1:**
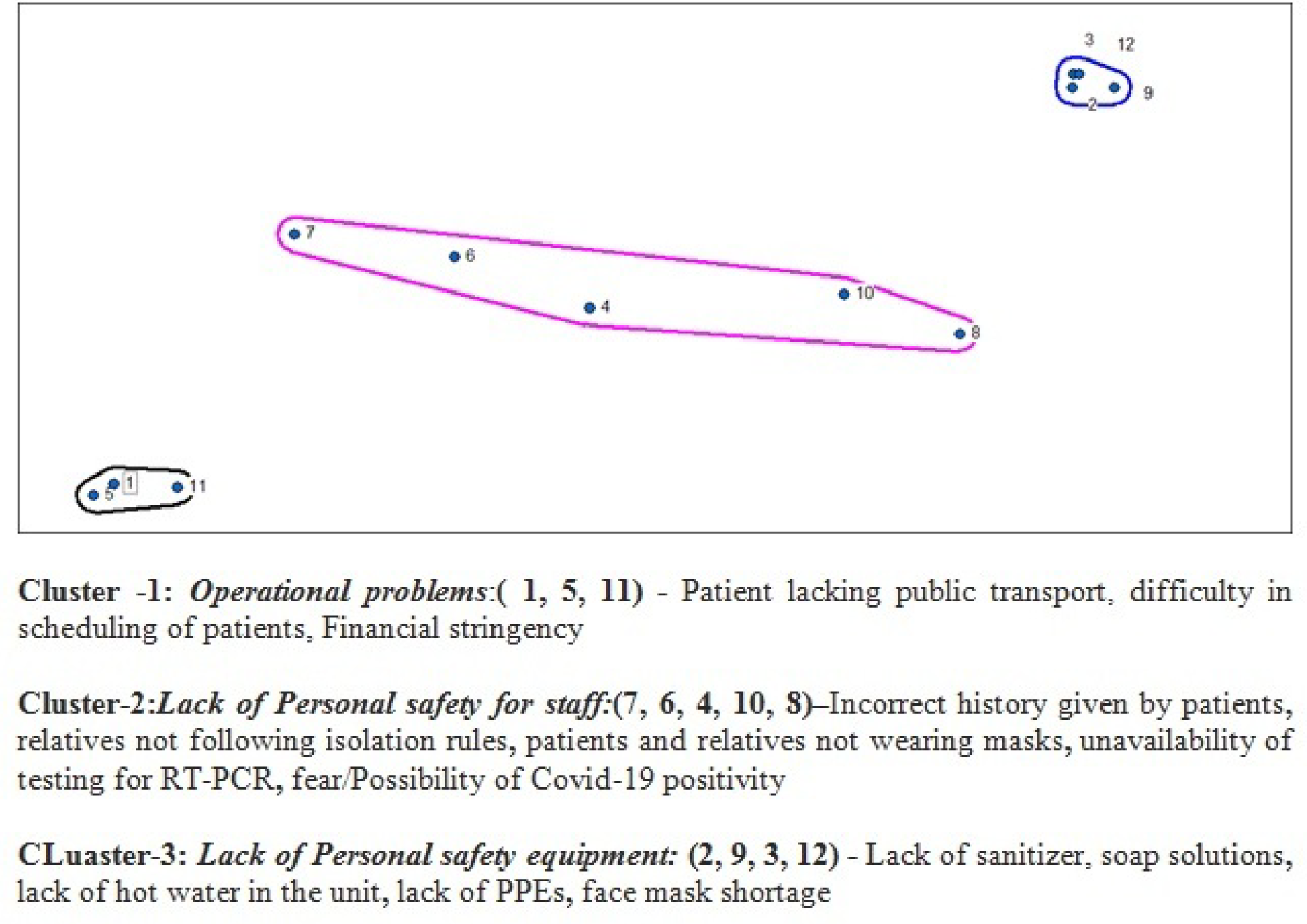
Cognitive domain of challenges faced by the staff (n=12)

**Figure 2.**
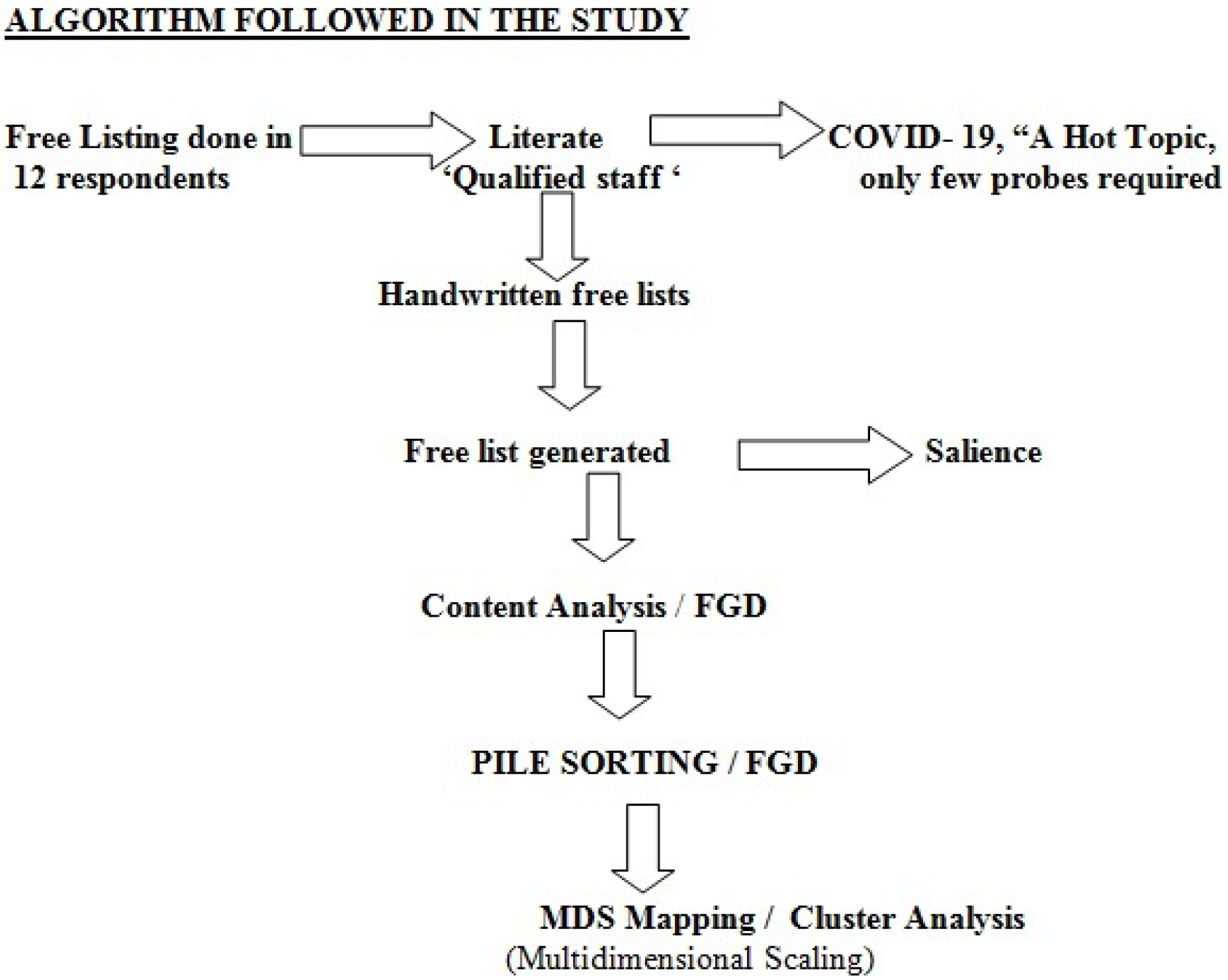

In prioritizing the different clusters, the first cluster chosen was the one dealing with shortages of the personal safety equipment for the dialysis staff. As a result of the information gathered from the dialysis staff, we immediately talked to the various stakeholders in the hierarchy of the institution and apprised them of our findings and showed them the results of the cluster mapping. They agreed to immediately order fresh standard masks, PPEs and sanitizers and provide hot water for the unit. The dialysis staffs were very appreciative of the immediate outcomes. There was remarkable transformation and positivity in the behavior and the work culture of the unit members.

The second group of clusters as regards the patients and their relatives not adhering to infection control practices, didactic meetings were held with them and the wearing of face masks and following isolation and infection control measures were stressed upon. The issues touched upon in the above two clusters were within our purview and we were able to address them successfully.

The problems regarding RT-PCR testing were for the hospital management to ponder upon, which they readily agreed to discuss and act upon. In fact, they did place the order for getting the RT-PCR machine.

The third cluster of issues pointed out was really beyond our control i.e. regarding the lack of public transport owing to lockdowns, disarray in the scheduling of patients (again due to lack of transport) and the general financial depression. This was under the domain of the Government and other official authorities. In fact, the patients petitioned the collector and the Government officials successfully to get them dropped to the hospital, using government ambulances.

Nowadays most people are usually more concerned about the RT-PCR testing and antibody testing, but our results showed that the concerns of dialysis staff were mainly centered upon protecting themselves (personal safety) by means of proper PPEs while caring for their patients.

The best strategy to prevent Covid-19 transmission is frequent hand washing, maintaining physical distancing, proper cough and sneeze etiquette, and regular environmental disinfection in the dialysis unit.^12,13,14^ Finding about the real issues bothering our health care staff was not a technical ability but was actually an elective, focused bonding towards them! The main direction of our outlook was to consider the staff working with us as an individual and not to just treat them like some stereotype. Finally above all the onus on us was to consider them as our fellow human beings facing fear of the unknown and uncertainity.^15^And not to consider in the least as some issue in cost accounting.^16^

The impact of the challenges of Covid-19 in a rural health care facility has its own multidimensional impact in the form of the lack of testing services, poor surveillance and acute shortage of equipment and PPEs.^17^Also the financial burden during these challenging times casts a shadow on the smooth functioning of the dialysis units directly impacting on patient care. The burnout rates are very high in challenging and high-pressure environments like busy dialysis units. The burden of the moral dilemmas afflicting the health care staff due to lack of equipment can easily lead to stress, depression, anxiety, Post-traumatic stress disorder and even suicides. Hence as a clinical director of a dialysis unit or as a dialysis manager it is our foremost duty to make sure the staff are provided with evidence based care and support materials.^18^Appreciation of the staff and expressing gratitude for their work during stressful times can raise their confidence levels and improve resilience.^19^

Health care workers constantly face an ethical dilemma which is fueled by the lack or inequitable distribution or even reusage of PPEs or masks.^20^The similar challenges faced by staff in some health care centers across Africa is of gargantuan proportions because of global jostling,^21^ lack of low cost face masks, and even lack of water for hand washing.^22^ Invaluable lessons were learnt at the time of Ebola in Africa and even in HIV control where dispelling myths and support to health care workers led the way forward.^23^

A recent qualitative study from China using semi structured in-depth interviews(empirical phenomenological approach) reported challenges faced by health care workers in covid-19 wards,exhaustion,fear of getting infected and infecting others(especially family).They suggest comprehensive support in form of adequate protective gear, effective communication, monitoring and surveillance of infection control.^24^

### Strengths and limitations

The study procedure was sequential in nature, where free list directed pile sort, which in turn offered direction to focus group, ultimately contributing the validity of the findings. The study was rapid and findings in visual format were easy to understand and evoke discussion among the participants. The findings were useful in understanding the dynamics for decision-making and action taking. Data saturation was achieved, adding more weightage to this study. There is scope for transferability of these findings to other departments in a hospital and various institutions in the surrounding areas of Pondicherry, all over India and elsewhere as such. The COREQ guidance (COnsolidated criteria for Reporting Qualitative research) as shown in Appendage 2 was applied while drafting this paper which added to the rigor, clarity and transparency of this study.^24,25,26^ However, the limitations of the present study should be kept in mind. The background of researcher and personal biases of the researcher could not be eliminated fully. Another limitation was that we did not further analyze the subgroups by separating the dialysis nurses, dialysis technicians and the doctors as our aim was to see the whole dialysis unit as one cohesive and patient centric group.

Despite all the challenges faced by the dialysis staff, the patients also have their challenges but yet they demonstrate remarkable resilience.^27^It would be worthwhile to get away from a traditional problem oriented approach to examine the strengths and vulnerabilities of our dialysis staff as done in patient oriented research.^28^

### Research Committee and Ethics clearance

The paper was cleared both by the research committee and by the Institutional Ethics committee vide: SMVMCH-EC-45/2020 −116/2020

### What is new in this study?

This is the first qualitative study in dialysis exploring the challenges faced by the dialysis unit staffs during the pandemic unleashed by the novel coronavirus SARS-COV-2!

A NLM-MESH/PUBMED ^29^ search of the databases did not yield any hits after putting the words used in the title of our study. According to us this is the first such study, which throws light on the perceptions and challenges faced or thought of by the dialysis health care workers during disasters and pandemics.

### Implications and outcomes of this study

This study also illustrates that such a qualitative research process by itself involves the subtle interrelationships and the intricacies of human interaction.^30^ We can use these research methods to resolve problems confronting our health care units.

This study throws new light on the thoughts, doubts, fears and priorities donning the minds of the dialysis staff members. This is also a novel and rapid way of getting more insight into the behaviors of the staffs working in dialysis units. It also will help in improving the group dynamics in a fast paced and high stress area of hemodialysis.

## Conclusion

The results of this qualitative study in dialysis staff show the significance of the thoughts; ideas and fears of the health care workers need to be addressed appropriately during these mitigating circumstances. This will lead to better and focused patient care, yielding better clinical outcomes in the dialysis units especially during pandemics and disasters.

## Supporting information

Supplemental file 1 COI disclosure form

supplemental Form 2 COI* disclosure form

Supplemental file3 Data Reporting Checklist

## Data Availability

All available data can be obtained by contacting the corresponding author. All of the individual participant data collected during the project is available, after de-identification, beginning 3 months and ending 5 years following article publication

## Acknowledgements

We thank the administration and the management of the Sri ManakulaVinayagar Medical College and Hospital, Pondicherry for permitting us to carry out this project.

## Disclosures

### Contributorship statement

RKP was involved in concept, planning, design of the study, collection of data, and proofreading and revision of the data.AD was involved in contributing to the concept, design, analysis, interpretation of the data, proofreading and revising the data and manuscript.

### Standard for Reporting

**COnsolidated criteria for reporting qualitative studies (COREQ** guidelines and Methodology was followed while writing this manuscript).

### Supplementary Material

Data Reporting checklist in PDF format.

