## Supplemental file3 Data Reporting Checklist for "Challenges Faced by Dialysis Unit Staff during COVID −19 times-A Qualitative Study"

### DATA REPORTING CHECK-LIST

**Reporting Guidelines:**

**COnsolidated criteria for reporting qualitative studies (COREQ): 32-item checklist was used in this study.**

No. / Item/ Guide questions/description

#### Domain 1: Research team and reflexivity

**Personal Characteristics**

1. **Interviewer/facilitator**: PRK conducted the interview or focus group discussions

2. **Credentials**: The Researcher PRK is an MD (Medicine), DNB (Nephrology) Clinical Fellowship in Adult Nephrology (University of Toronto), ExWHO Consultant, Republic of Seychelles, East Africa

Prof.AD is an MD (Community Medicine) MPS, MHPE, FAIMER Fellow (2008) WHO in-country Fellow (2011), IFME (2015)

3. **Occupation**: PRK is the Senior Consultant Nephrologist and HOD at the Sri Manakula Vinayagar Medical College and Hospital, Puducherry, India. Prof AD is HOD of community Medicine at the Sri Manakula Vinayagar Medical College and Hospital, Puducherry India.

4. Gender: Both are Males

5. **Experience and training**: PRK has undergone training in Qualitative research and is a member of the research committee and the IEC at the Sri Manakula Vinayagar Medical College and Hospital, Puducherry, India He also ran a CQI project in vascular access in Nephrology for over two years at the Sunny brook Health Sciences centre, University of Toronto, Canada. Prof AD is the pioneer of the Qualitative research in India. He holds yearly Qualitative research courses as a Lead Faculty/Facilitator since last 12 years. He is the Dean of Research at the Sri Manakula Vinayagar Medical College and Hospital, Puducherry, India.

**Relationship with participants**

6. Relationship established: A relationship was established prior to study commencement.

7. Participant knowledge of the interviewer

What did the participants know about the researcher? There was deep concern and feelings about the challenging a time working in the dialysis unit during the Covid -19 pandemic. And any project or studies to help bring out these concerns were important in the participant’s perspective. The researcher was in daily interaction with the unit members so there was already rapport between both the respondent and the interviewer.

8. Interviewer characteristics: The objective of the study was a very important issue affecting the morale of the unit. The whole focus was on the outcome of this project safety for both the staff and the patients. Personal bias could not be eliminated totally. The interviewer as a clinician-researcher was keen to help improve the mindset of the dialysis staff. The reason for conducting the study was to dispel the morbid fears and to understand the deficiencies, inadequacies and defects in the dialysis unit at the time of the covid19 crisis.

#### Domain 2: study design

Theoretical framework

9. **Methodological orientation and Theory**

Content Analysis was used to underpin the study. Concept mapping gave the structural framework for the study.

**Participant selection**

10. Sampling: Participants were selected by purposive sampling.

11. **Method of approach**: Participants were approached sitting in a circular group for the interview/FGD and then again individually face-to-face while pile sorting was done.
12. **Sample size**: There were 12 participants in the study.
13**. Non-participation**: No one refused to participate.

**Setting**

14. **Setting of data collection**: The data was collected at the dialysis unit in Sri Manakula Vinayagar Medical College and Hospital, Puducherry, India.
15. **Presence of non-participants**: None
16. **Description of sample**: The important characteristics: The 12 participants comprised of five nurse, five dialysis Technicians and two doctors.They are reported as such in the paper.

Date of the study was 18thJuly, 2020

**Data collection**

17. Interview guide: The questions, prompts, were provided by the authors. However no pilot testing was done.
18. Repeat interviews: Repeat interviews carried out once before pile sorting was done.
19. Audio/visual recording Paper transcripts were done in the study.
20. Field notes Field notes were made during and after the interview / focus group discussions.
21. Duration The duration of the interviews or focus group was 10-15 minutes.
22. Data saturation Data saturation was not discussed.
23. Transcripts returned Transcripts were returned to participants for comment and corrections. Participants actually checked their own transcripts for greater accuracy.

**Domain 3: analysis and findings**

**Data analysis**

24. Number of data coders: Two
25. Description of the coding tree: Did authors provide a description of the coding tree? No, as a computer soft ware was used.
26. Derivation of themes: The Themes were derived from the data.
27. Software was used to manage the data: Anthropac 4.983/X (By Stephen P Borgatti copyright1992, Analytic Technologies) was used for free listing, pile sorting and to create the cluster mapping to finally show the digital visualization.
28. Participant checking: Yes the Participants provided feedback on the findings.

**Reporting**

29. Quotations presented: Participant quotations were presented to illustrate the themes / findings Each quotation was identified.
30. Data and findings consistent: There was consistency between the data presented and the findings.
31. Clarity of major themes: The major themes were clearly presented in the findings.
32. Clarity of minor themes: There is a description of diverse cases and minor themes were also discussed.
